# Adherence to International Pharmacogenomic Recommendations in Paediatric Cancer Care: A Cohort Analysis Embedded Within the MARVEL-PIC Randomised Trial

**DOI:** 10.64898/2026.04.15.26348678

**Authors:** Aniket Chawla, Sean Carter, Roxanne Dyas, Elizabeth Williams, Claire Moore, Rachel Conyers

## Abstract

**Background:** Pharmacogenomic testing (PGx) can optimise drug efficacy and minimise toxicity, but the extent of prescriber adherence to PGx recommendations remains unclear. We aimed to quantify clinician adherence to international genotype-guided prescribing recommendations in a cohort of paediatric oncology patients.

**Methods:** We reviewed files of children enrolled in the MARVEL-PIC (NCT05667766) randomised control trial, who had PGx recommendations available. Patients were included if 12 weeks had passed since their PGx report was released to clinicians. Prescribing events were identified for actionable PGx recommendations, and classified as “explicitly followed”, “inadvertently followed”, or “not followed”. Adherence was assessed by patient, drug, and recommendation.

**Results:** 2,063 PGx recommendations were available for 216 patients. 64 (3.1%) recommendations were actionable for 44 patients and 10 drugs within the 12-week study period. Recommendations were explicitly followed in 57/288 (19.8%) of prescribing events, inadvertently followed in 145 (50.3%), and not followed in 86 (29.9%). Mercaptopurine demonstrated the highest rate of explicit adherence (87.5%). No significant associations were observed between adherence and age group, cancer type, drug type, or strength of recommendation.

**Conclusion:** Adherence to pharmacogenomic recommendations was very low, highlighting the need to understand barriers to PGx implementation, and consideration of clinical decision supports to facilitate adherence.

**Plain Language Summary:** Pharmacogenomic medicine (PGx) looks at how our genes affect our response to drugs, including their effectiveness and toxicity. Through genetic analysis we can create recommendations for drug dosing, avoidance, and monitoring. The MARVEL-PIC study aims to understand if having PGx recommendations decreases the rate of adverse events in children with cancer.

We aimed to understand how often prescribers follow PGx recommendations after they are made available, in the MARVEL-PIC trial. To do this, we reviewed medical records and identified relevant prescribing events. We marked these as “recommendation explicitly followed”, “recommendation not followed”, or “recommendation inadvertently followed” (where the recommendation was followed, but it wasn’t clear if this due to PGx).

We found that when recommendations were available, they were only explicitly followed in around 20% of cases. In 50% of cases, they were followed but it was unclear whether this was due to PGx. In the remaining 30%, they were not followed. We also found that alerts on our electronic system were fired in about 80% of events where the recommendation was not followed, but did not change the outcome. These findings show that prescriber adherence to PGx recommendations is low. We need to better understand why this is the case and implement more specific tools to assist prescribers in following recommendations.

**Article Highlights:**

- Pharmacogenomic (PGx) testing can reduce adverse drug reactions by guiding drug choice, dosing, and monitoring.
- !Prescriber to PGx recommendation adherence has not been widely investigated.
- Retrospective analysis showed that explicit adherence to recommendations occurred in only 19.8% of relevant prescribing events.
- In 50.1% of prescribing events, recommendations were followed, but there was no clear reference to PGx.
- Mercaptopurine had the highest explicit adherence (87.5%) from the drugs analysed.
- There were no statistically significant associations between adherence and age group, cancer type, drug type, or recommendation strength.
- Recommendations were explicitly followed in 29% of events where an interruptive alert was fired, and inadvertently followed in 8%.
- Tailored interruptive alerts have been shown to increase adherence in other studies, suggesting that the specific design of interruptive alerts may influence adherence.
- We concluded that explicit prescriber adherence to PGx recommendations is very low (19.8%), and further research needs to be done to understand barriers to implementation.

## 1.0 Introduction

Pharmacogenomics (PGx) is a field of precision medicine that studies how genotypic variation contributes to the toxicity and efficacy of medications [1]. Genotype-informed prescribing has the potential to reduce adverse drug reactions (ADRs). The utility of PGx at scale was demonstrated in a landmark 2023 study of 6,944 European adults, which showed that genotype-guided prescribing reduced ADRs by 30% (OR 0.70 [95% CI 0.54–0.91]; p=0.0075) [2]. There is also the potential for health resource reduction; a 2016 meta-analysis demonstrated robust evidence of cost-effectiveness of genotyping prior to treatment with a number of drugs [3]. However, in paediatrics neither the utility nor cost-effectiveness of PGx had been proven; this forms the basis of the Minimising Adverse Drug Reactions and Verifying Economic Legitimacy: Pharmacogenomic Implementation in Children trial (MARVEL-PIC; NCT05667766) [4].

International consortia, such as the Dutch Pharmacogenetics Working Group (DPWG) and the Clinical Pharmacogenetics Implementation Consortium (CPIC) have provided evidence-based frameworks for genotype informed prescribing. To date CPIC has included 573 gene-drug pairs of which 100 have Tier A recommendations available (i.e. where the evidence includes consistent results from well-designed, well-conducted studies) [5]. Despite recommendations across many supportive care and cancer medications, widely conducted PGx testing in paediatric oncology is limited to variants of *TPMT* and *NUDT15* in children with acute lymphoblastic leukaemia [6].

The MARVEL-PIC randomised control trial (RCT) is recruiting children with a new cancer diagnosis across four states in Australia to assess the impact of pre-emptive pharmacogenomic testing on incidence of ADRs in children with cancer [4]. Children in both the intervention and control arms undergo pharmacogenomic testing for variations in 14 different genes, with prescribing implications for 32 drugs (Table 1). The primary oncology physician is provided with prescribing recommendations based on a patient’s genotype when available (intervention arm), or after 12 weeks (control arm). Enactment of recommendations is left to the discretion of the treating clinical team, with no interference from the study team. Incidence of ADRs is then noted during the first 12 weeks at three separate time points, and comparison made between the intervention arm (where PGx recommendations were available) and control arm (where PGx recommendations were not).

**Table 1:**
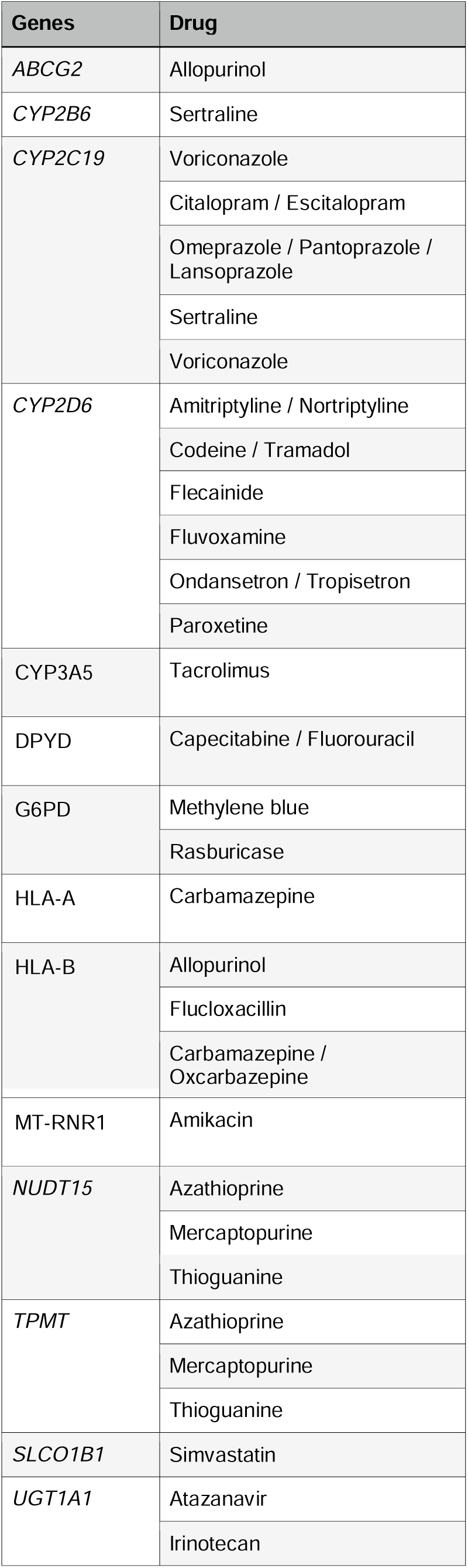
Gene-drug pairs assessed in the MARVEL-PIC study.

Adherence to international PGx recommendations in paediatric oncology cohorts has not been widely quantified in the literature. This study aimed to determine prescriber adherence to international pharmacogenomic recommendations for 32 gene-drug pairs investigated in the MARVEL-PIC cohort recruited thus far, through a retrospective chart review.

## 2.0 Methods

### 2.1 Ethics

This study was conducted as a sub-study of the MARVEL-PIC trial, and ethical approval was covered under the parent MARVEL-PIC human research ethics approval, obtained from the Royal Children’s Hospital Ethics Committee (HREC/89083/RCHM-2022).

### 2.2 Study Population

Patients included in this study were required to be enrolled in the MARVEL-PIC RCT (intervention or control arm) between March 2023 and September 2025, have had a PGx report available for 12 weeks, and have genotype reports with prescribing recommendations available in their Electronic Medical Record (EMR).

### 2.3 Data Collection

Demographic data collected from the MARVEL-PIC study included age at diagnosis, sex at birth, cancer diagnosis, and pharmacogenomic recommendations based on genotype/phenotypic metaboliser status. A retrospective chart review of the EMR was performed to identify relevant prescribing opportunities for each patient, based on their individual PGx report.

### 2.4 Categorisation of PGx reporting, Prescribing Events, and Adherence

Duplicate recommendations for drugs of the same class, indication, and genetic metabolism pathway were excluded, as these drugs were not likely to be prescribed in tandem. This included recommendations for amitriptyline/nortriptyline, capecitabine/fluorouracil, citalopram/escitalopram, codeine/tramadol, omeprazole/pantoprazole, and ondansetron/tropisetron.

The remaining patient PGx report recommendations were coded by a single researcher as “actionable”, “not actionable”, or “standard prescribing” (Figure 1). Actionable recommendations were defined as those requiring changes to dosing, biochemical/therapeutic drug monitoring (TDM), or avoidance of a drug. Recommendations were only deemed “actionable” if the patient was prescribed that drug during the study period, or if, where a drug was recommended to be avoided, a suitable alternative drug was prescribed for the relevant indication during the study period. “Not actionable” recommendations included those where a recommendation was present, but there was no opportunity to follow it during the prescribing period (i.e. drug/alternative drug for same indication not prescribed). “Standard prescribing” was defined as where the recommendation was to initiate standard-of-care dosing.

**Figure 1:**
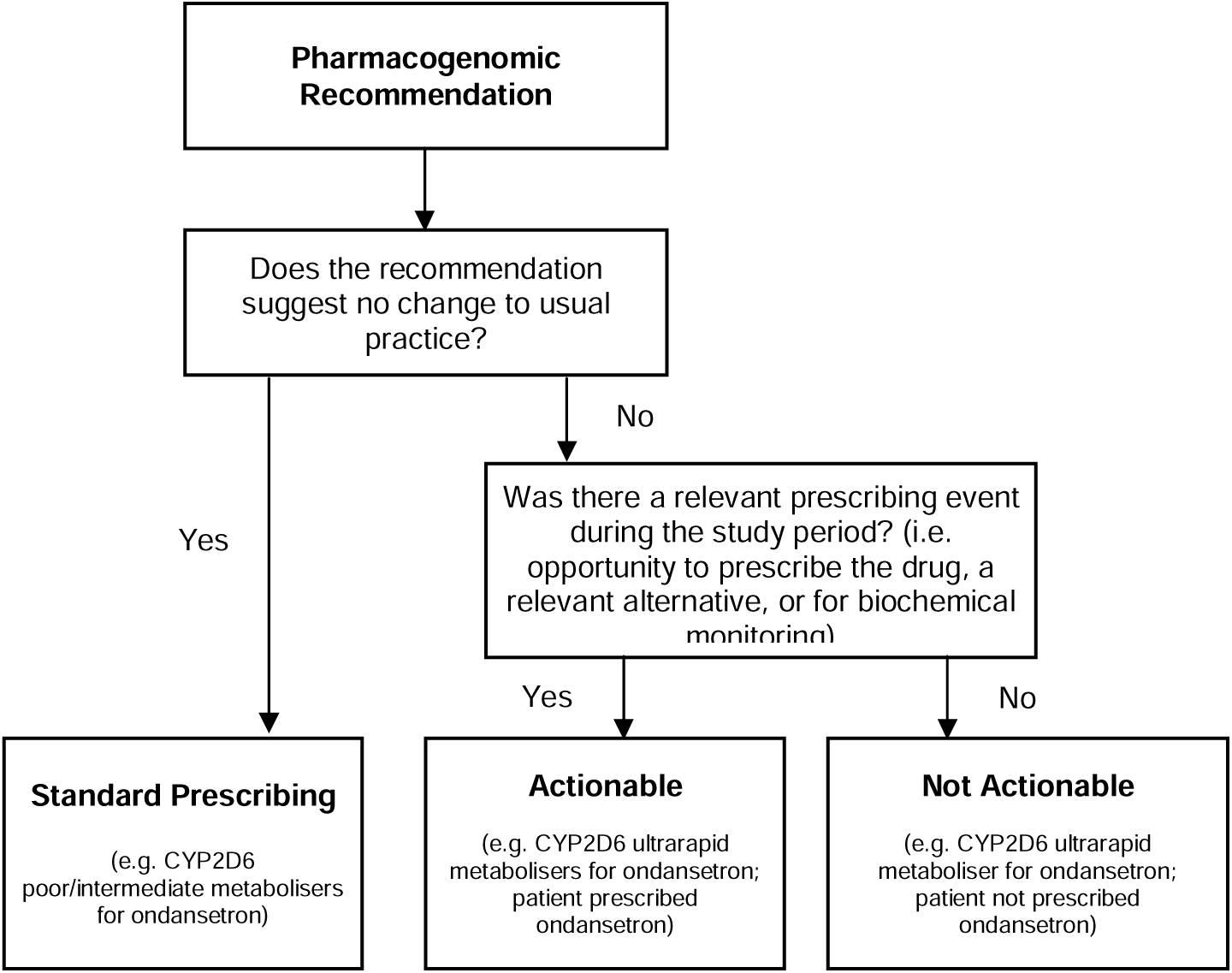
Flow chart demonstrating classification of recommendations

For each actionable recommendation, we recorded all prescribing events within the 12 weeks following report release. A prescribing event was defined as any prescription of the relevant drug, or, where avoidance was recommended, prescription of a suitable alternative for the same clinical indication. For recommendations requiring increased TDM, a prescribing event was defined as each instance of TDM performed.

Each prescribing event was categorised as “explicitly followed”, “not followed”, or “inadvertently followed” according to whether the relevant PGx recommendation was followed. (Table 2) “Inadvertently followed” events were defined as those where a recommendation was followed, but there was no clear evidence of this prescribing event being influenced by pharmacogenomic recommendations.

**Table 2:** Relevant terms and definitions.

| Term | Definition |
| --- | --- |
| Prescribing event | Any instance in which a PGx recommendation could have been followed (e.g., the relevant drug prescribed; alternative drug prescribed for the same indication; or biochemical/therapeutic drug monitoring performed). |
| Standard prescribing | The PGx recommendation does not deviate from standard practice (e.g. "Use standard dosing") |
| Actionable | A specific action is recommended (e.g. dose change, monitoring, or avoidance of a drug) AND this recommendation is applicable during the study period, i.e. the drug has been prescribed, or if avoidance is recommended, an alternative drug was prescribed for the same indication. |
| Not Actionable | A specific action is recommended, but it is not applicable during the study period, i.e. drug not prescribed, or no alternative drug prescribed for same indication. |
| Explicitly Followed | The recommendation was implemented and documentation explicitly attributes this to PGx recommendation. |
| Inadvertently Followed | The recommendation was implemented, but there is no documentation linking the action to PGx (e.g., a "avoid codeine/tramadol" recommendation is met because an alternative opioid was prescribed, without reference to PGx). |
| Explicit Adherence | Proportion of recommendations implemented with explicit documentation attributing the action to PGx, i.e. "Explicitly Followed" / "Total" |
| Overall Adherence | Proportion of recommendations implemented regardless of documentation, i.e., ("Explicitly Followed" + "Inadvertently Followed") / "Total" |

After categorising prescribing events, adherence was graded as high (≥75%), moderate (50–74.9%), low (25–49.9%), or very low (<25%) for each patient, drug, and recommendation. Adherence was assessed using two measures: (1) explicit adherence (“explicitly followed” prescribing events) and (2) overall which included both “explicitly followed” and “inadvertently followed” prescribing events.

Interruptive alerts were incorporated into the EMR as part of the MARVEL-PIC study; when a drug-gene interaction was detected, the prescriber was alerted to refer to the full PGx report. The recommendation itself was displayed in the interruptive alert. Prescribing events were correlated with a record of interruptive alerts fired on the EMR to determine whether an alert was fired in each event. Interruptive alerts were not fired in prescribing events where the corresponding recommendation was to avoid a drug and prescribe a suitable alternative, or for biochemical/therapeutic drug monitoring.

### 2.5 Statistical Analysis

Descriptive statistics were calculated. Fisher-Freeman-Halton Test was performed due to low contingency table cell counts, to assess the relationship between adherence and age, disease group, drug type (chemotherapy vs treatment vs supportive), and strength of recommendation (as determined by DPWG/CPIC). p<0.05 was deemed to be significant. Cramér’s V values were manually calculated to describe effect size of any associations. The statistical software used was R Commander.

## 3.0 Results

### 3.1 Study Population

Between March 2023 and September 2025, 245 children aged 0-18 years received pharmacogenomic testing as part of the MARVEL-PIC study, at the Royal Children’s Hospital in Melbourne, Australia. Patients for whom 12 weeks had lapsed since release of their report (n=216), were included.

### 3.2 Baseline Characteristics

120 patients (55.6%) were male and 96 (44.4%) were female. The median age at diagnosis was 6.5 years (range 0.2-17.3 years). 91 patients (42.7%) had haematological malignancies, 82 (38.5%) had solid malignancies, 17 (8.0%) had CNS malignancies, and 23 (10.8%) patients received stem cell transplants. Clinical characteristics of all patients and included patients are listed in Table 3.

**Table 3:** Characteristics of study population.

|  | All patients<br>(n=216) | Included<br>patients (n=44) |
| --- | --- | --- |
| **Age at Diagnosis** |  |  |
| Mean | 7.54 | 8.24 |
| SD | 5.29 | 5.16 |
| Median | 6.5 | 8.1 |
| Range | 0.2-17.3 | 0.2-17.3 |
| **Age Group** |  |  |
| 0-4.9 | 92 (42.6%) | 15 (34.1%) |
| 5-9.9 | 44 (20.4%) | 11 (25.0%) |
| >10 | 80 (37.0%) | 18 (40.9%) |
| **Sex at birth** |  |  |
| Male | 120 (55.6%) | 32 (72.7%) |
| Female | 96 (44.4%) | 12 (27.3%) |
| **Diagnostic group** |  |  |
| CNS | 17 (8.0%) | 4 (9.1%) |
| Solid | 82 (38.5%) | 12 (27.8%) |
| Haematological | 91 (42.7%) | 21 (47.7%) |
| Transplant | 23 (10.8%) | 7 (15.9%) |

### 3.3 PGx Reporting and Prescribing Events

2,063 individual gene-drug prescribing recommendations (median 9, range 0-22) were made for these 216 patients. 510 (24.9%) recommendations were excluded as duplicates. 357 (17.4%) recommendations were deemed “standard prescribing”. Of the remaining 1,196 recommendations, 1,132 were not actionable (i.e. did not have an associated prescribing event during the 12-week period). 64 (3.1%) recommendations for 10 drugs were deemed actionable, for a total of 44 patients (Figure 2). 288 prescribing events were identified for analysis, for these 44 patients. Of these, 205 (71.2%) prescribing events occurred in an inpatient setting, 44 (15.3%) in an outpatient setting, and 39 (13.5%) at discharge from hospital.

**Figure 2:**
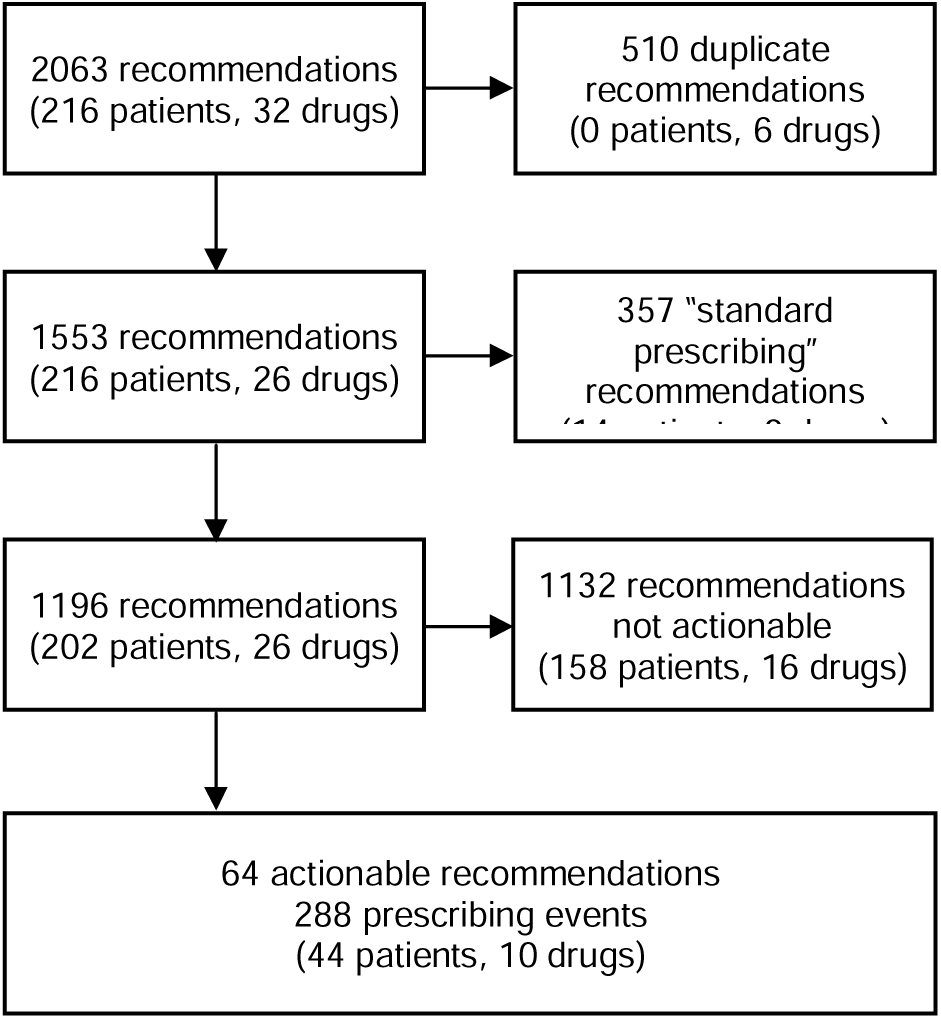
Flow chart demonstrating the number of actionable recommendations relevant within the 12-week study period

### 3.4 Adherence

In 57/288 (19.8%) of identified prescribing events, the PGx recommendation was explicitly followed, while in 86/288 (29.9%), it was not followed. In the remaining 145 prescribing events (50.3%), the recommendation was inadvertently followed. Examples of a recommendation being inadvertently followed include where a patient was recommended to avoid codeine/tramadol, and a non-codeine/tramadol opioid was prescribed, but without reference to PGx. Thus, the rate of explicit was 19.8% (57 events), and overall adherence was 70.1% (202 events).

Of the 44 patients analysed, 16 (36.4%) had at least one PGx recommendation explicitly followed during at least one prescribing event. However, 29 (65.9%) patients demonstrated very low rates (<25%) of explicit adherence. Only seven (15.9%) demonstrated high adherence (≥75%). When considering overall adherence (i.e. including prescribing events where a recommendation was followed without explicit reference to pharmacogenomics), 24 (54.5%) of patients demonstrated high overall adherence and 10 (22.7%) very low overall adherence. These findings are illustrated in further detail in Table 4.

**Table 4:** Recommendation adherence rates by patient, drug, and recommendation.

|  |  | Very Low (<25%) | Low (25-50%) | Moderate (50-75%) | High (≥75%) | Total |
| --- | --- | --- | --- | --- | --- | --- |
| **By patient** |  |  |  |  |  |  |
|  | Explicit adherence | 29 (65.9%) | 3 (6.8%) | 5 (11.4%) | 7 (15.9%) | 44 (100%) |
|  | Overall adherence | 10 (22.7%) | 2 (4.5%) | 8 (18.2%) | 24 (54.5%) | 44 (100%) |
| **By drug (e.g. allopurinol)** |  |  |  |  |  |  |
|  | Explicit adherence | 6 (60%) | 2 (20%) | 1 (10%) | 1 (10%) | 10 (100%) |
|  | Overall adherence | 2 (20%) | 1 (10%) | 1 (10%) | 6 (60%) | 10 (100%) |
| **By individual recommendation (see Supplementary Table 1)** |  |  |  |  |  |  |
|  | Explicit adherence | 10 (58.8%) | 2 (11.8%) | 1 (5.9%) | 4 (23.5%) | 17 (100%) |
|  | Overall adherence | 3 (17.6%) | 2 (11.8%) | 1 (5.9%) | 11 (64.7%) | 17 (100%) |

Mercaptopurine was the only drug with a high rate (87.5%) of explicit pharmacogenomic adherence (Table 5). Recommendations for thioguanine were explicitly followed in one of two total (50%) prescribing events. Voriconazole, ondansetron/tropisetron, and omeprazole/pantoprazole also demonstrated above-zero rates of explicit adherence (41.4%, 30.4%, and 22.0%, respectively), whereas recommendations for amitriptyline/nortriptyline, carbamazepine, codeine/tramadol, and sertraline were not adhered to in any prescribing event. 6/10 (60%) of prescribed drugs demonstrated high overall adherence of recommendations.

**Table 5:** Recommendation adherence rates and grades, by drug.

|  |  | n<br>(patients) | n<br>(events) | Rate<br>(events) | Grade |
| --- | --- | --- | --- | --- | --- |
| **Recommendation explicitly followed** |  |  |  |  |  |
|  | Amitriptyline | 9 | 36 | 0.0 | Very low |
|  | Amitriptyline / Nortriptyline | 2 | 12 | 0.0 | Very low |
|  | Carbamazepine | 1 | 4 | 0.0 | Very low |
|  | Codeine / Tramadol | 12 | 73 | 0.0 | Very low |
|  | Mercaptopurine | 5 | 16 | 87.5 | High |
|  | Omeprazole /<br>Pantoprazole | 19 | 41 | 22.0 | Very low |
|  | Ondansetron / Tropisetron | 6 | 69 | 30.4 | Low |
|  | Sertraline | 1 | 5 | 0.0 | Very low |
|  | Thioguanine | 1 | 2 | 50.0 | Moderate |
|  | Voriconazole | 8 | 29 | 41.4 | Low |
| **Recommendation inadvertently followed** |  |  |  |  |  |
|  | Amitriptyline | 9 | 36 | 100.0 | High |
|  | Amitriptyline / Nortriptyline | 2 | 12 | 100.0 | High |
|  | Carbamazepine | 1 | 4 | 100.0 | High |
|  | Codeine / Tramadol | 12 | 73 | 100.0 | High |
|  | Mercaptopurine | 6 | 16 | 87.5 | High |
|  | Omeprazole /<br>Pantoprazole | 19 | 41 | 22.0 | Very low |
|  | Ondansetron / Tropisetron | 6 | 69 | 34.8 | Low |
|  | Sertraline | 1 | 5 | 0.0 | Very low |
|  | Thioguanine | 1 | 2 | 50.0 | Moderate |
|  | Voriconazole | 8 | 29 | 100.0 | High |

Four specific recommendations demonstrated high levels of explicit adherence. Three of these recommendations were aimed at reducing dosing of mercaptopurine (Recommendations 31-33, adherence 83.3-100%; see Supplementary Table 1 for comprehensive list of recommendations), and one at therapeutic drug monitoring of voriconazole (Recommendation 60, adherence 80%). Further specific adherence rates by recommendation can be found in Supplementary Table 2.

Interruptive alerts on the Electronic Medical Record (EMR) were fired for 100 prescribing events. The recommendation was explicitly followed in 29 (29.0%) of events, inadvertently followed in eight (8.0%), and not followed in 63 (63.0%).

No statistically significant associations (p<0.05) were found between adherence and age group, type of cancer, type of drug, or strength of recommendation (Supplementary Table 3).

## 4.0 Discussion

The implementation of pharmacogenomic-guided prescribing into clinical practice remains challenging. We aimed to understand the adherence rates to specific international genotype-informed prescribing recommendations in the MARVEL-PIC RCT cohort [4]. Within our cohort, only 19.8% of prescribing recommendations were explicitly followed. 65.9% of the analysed patients displayed a very low rate (<25%) rate of adherence, and only recommendations for mercaptopurine and thioguanine demonstrated explicit adherence rates of ≥50%. Four individual recommendations were highly explicitly adhered to (≥75% adherence). Three of these were related to lower dosing of mercaptopurine (Recommendations 31-33), and one recommended meticulous therapeutic drug monitoring of voriconazole levels (Recommendation 60). When considering prescribing events where recommendations were inadvertently, adherence rate increased from 19.8% (explicit adherence) to 72.5% (overall adherence).

Of 86 prescribing events in which pharmacogenomic recommendations were not followed, reasons for not following the recommendation were documented in only two. In both of these instances, the clinician expressed confusion regarding the pharmacogenomic advice and ultimately did not follow it. In the remaining 84 events, no reason was documented for not following the recommendation. As documentation does not always capture the intricacies of clinical decision-making, there may have been prescribing events where recommendations were considered but deemed clinically inappropriate, but these were not documented.

In 50.3% of prescribing events, our recommendations were “inadvertently followed”. The most common recommendations to be inadvertently followed were Recommendation 8 and Recommendation 22, which recommended to avoid codeine/tramadol and amitriptyline, respectively. The high incidence of this is likely a reflection of local prescribing patterns; at our centre, oxycodone is the first-line opioid of choice, and gabapentin is the first-line drug to treat neuropathic pain. As such, these recommendations were likely to have been followed regardless of pharmacogenomic advice, and cannot be considered as true adherence to recommendations. There may also have been prescribing events where these recommendations were acknowledged and guided the prescription, but this was not documented.

Our findings closely mirror those of Tremmel et al., who found that 21.5% (55/255) patients had actionable recommendations, compared to 20.8% in the present study. They noted that overall accordance to pharmacogenomic guidelines was 75.5%, broadly similar to our overall adherence rate of 70.1%. However, direct comparison is limited by differences in terminology and methodology differences; Tremmel et al. considered “recommendations” while we considered “prescribing events”. Furthermore, Tremmel et al. assessed an adult cohort, whereas we assessed a paediatric cohort. Notably, they reported 100% accordance to recommendations regarding chemotherapeutic drugs (5-fluorouracil, capecitabine, mercaptopurine, and irinotecan). Our overall adherence for chemotherapy drugs was lower -- 14/16 (87.5%) events for mercaptopurine and 1/2 (50.0%) events for thioguanine. Undocumented clinical judgement and the small number of evaluable patients may explain this difference. Tremmel et al. also characterised recommendations as “pre-emptive”, where PGx recommendations were available prior to prescription, and “reactive”, where recommendations were generated after prescription, demonstrating 100% and 30% enactment in these categories, respectively. One significant strength of their study was that, as their cohort consisted of adults, a wider variety of drugs with available recommendations was prescribed as compared to our paediatric cohort.

The use of interruptive alerts to increase pharmacogenomic adherence has also been discussed in the literature. Nguyen et al. reported a 92% adherence rate to pharmacogenomic recommendations, attributed to their use of specific interruptive alerts [8]. A 2016 implementation study found that interruptive alerts significantly reduced the prescription of codeine to patients with high-risk CYP2D6 phenotypes. In this study, only one patient out of 53 with high-risk phenotypes (“possible ultrarapid metaboliser”) was prescribed codeine, compared to 173/543 (32%) without high-risk phenotypes [9]. Notably, the one patient who was prescribed codeine, had previously tolerated codeine well. Similarly, Bell et al. demonstrated that interruptive alerts incorporating specific action pathways (e.g. cancel, modify dose, continue) guided 95% of relevant prescribing decisions, encouraging genetic testing before prescription of thiopurines in 14/15 patients and avoidance of codeine in 5/6 patients [10]. All three of these studies emphasise the importance of clinical decision support through interruptive alerts, with Nguyen et al. also noting that co-designing interruptive alerts with clinicians also likely contributed to their high adherence rate [8].

While our study also utilised interruptive alerts, these were less specific, only encouraging prescribers to review the pharmacogenomic recommendations rather than listing the recommendation itself. As such, in 63/100 (63.0%) prescribing events where an interruptive alert was fired, the recommendation was not followed. Making pharmacogenomic recommendations more easily accessible to prescribers, with prescriber input into implementation, thus may also help increase adherence.

Alongside the retrospective nature of our data on adherence, and less specific interruptive alerts, low adherence in our cohort may have occurred due to limited awareness of pharmacogenomic medicine, and limited awareness of its proven benefits in patient care. This has been demonstrated previously in the literature, with a qualitative analysis of healthcare professional prospectives highlighting lack of knowledge, inadequate guidance, and low confidence in prescribers’ ability to implement pharmacogenomic data into practice, as key barriers to the adoption of pharmacogenomic medicine within Australia [11]. Beyond this study, literature around medical practitioners’ attitudes towards pharmacogenomic prescribing remains sparse, with other reviews primarily focusing on attitudes around pharmacogenomic testing.

Our study had a number of strengths and limitations. Data collection and analysis reflected real-world prescribing nuances, with prescribing events being classified as “explicitly followed”, “not followed”, or “inadvertently followed”, and analysed against two metrics (“explicit adherence” and “overall adherence”). Furthermore, our analysis moves beyond binary outcomes to demonstrate that adherence may be inconsistent even for the same patient; a single instance of a recommendation being followed by one prescriber does not guarantee uniform adoption by others. Our study is also one of few to quantitively investigate adherence to pharmacogenomic recommendations, and unique in its specificity to a paediatric oncology cohort. However, we were limited by a small sample size of actionable recommendations. This is primarily as many of the drugs that we provided recommendations for (e.g. sertraline, simvastatin, flecainide), were not prescribed during the 12-week study period for this paediatric cohort. Furthermore, a number of recommendations (e.g. to avoid codeine/tramadol and amitriptyline) did not significantly depart from Australian local practice, and thus true adherence was difficult to analyse. The retrospective nature of this study, as well as our data collection method from our EMR was also limiting, as we were unable to determine why recommendations were not followed, or whether they were considered during the prescribing process.

Despite these limitations, we have demonstrated significant findings of poor clinician adherence to pharmacogenomic recommendations. Qualitative investigations into clinician attitudes towards pharmacogenomic medicine are recommended to identify barriers to implementation, with educational interventions being tailored to address these barriers. We also recommend further quantitative analysis into adherence, with adherence data collected in real time, large sample sizes and a wider range of drugs that are more likely to be prescribed to children. To increase adherence in the short-term period, more tailored interruptive alerts on the EMR are suggested.

## 5.0 Conclusion

Our single-site retrospective review demonstrates that explicit adherence to pharmacogenomic recommendations is very low (19.8%) in children with cancer. This highlights a substantial gap in implementation of pharmacogenomic medicine, even when recommendations are available to clinicians. Due to limitations in our study design, we were unable to assess the reasoning behind low adherence rates. However, we identified accessibility and education as key targets to improving adherence. Future studies with larger cohorts, a broader range of medications more specific to paediatric medicine, and qualitative analysis into pharmacogenomic decision-making, are necessary to increase the uptake and utility of pharmacogenomic-guided prescribing in children.

## Supporting information

Supplementary Material S1: Recommendations by Drug

Supplementary Table S2: Prescribing events and adherence rates/grades by individual PGx recommendation

## Author Contributions

AC: conceptualisation, methodology, data curation, investigation, formal analysis, project administration, writing – original draft

SC: conceptualisation, methodology, writing – review and editing

RD: conceptualisation, writing – review and editing

EW: conceptualisation, writing – review and editing

CM*: conceptualisation, methodology, supervision, writing – review and editing

RC*: conceptualisation, methodology, supervision, writing – review and editing

* Both authors contributed equally and are co-senior authors of this manuscript.

## Disclosures

### Funding Statement

MARVEL-PIC is funded through a nationally competitive Medical Research Future Fund Grant (Genomics Health Future Fund MRF/2024900). This present study did not require any additional funding.

### Conflicts of Interest

All authors declare no conflicts of interest associated with this study.

## Abbreviations

ADR: Adverse Drug Reactions
CPIC: Clinical Pharmacogenetics Implementation Consortium
CNS: Central Nervous System
DPWG: Dutch Pharmacogenetics Working Group
EMR: Electronic Medical Record
PGx: Pharmacogenomics
MARVEL-PIC: Minimising Adverse Drug Reactions and Verifying Economic Legitimacy: Pharmacogenomic Implementation in Children
RCT: Randomised Control Trial
TDM: Therapeutic Drug Monitoring

## Data Availability

Deidentified datasets generated during and/or analysed during the current study are available from the corresponding author upon reasonable request.

## Notes

### Competing Interest Statement

The authors have declared no competing interest.

### Clinical Trial

NCT05667766

