## Supplementary Material S1: Recommendations by Drug for "Adherence to International Pharmacogenomic Recommendations in Paediatric Cancer Care: A Cohort Analysis Embedded Within the MARVEL-PIC Randomised Trial"

**Allopurinol**

| **Gene** | **Genotype** | **#** | **Recommendation** | **Classification** | **Potentially actionable?** |
| --- | --- | --- | --- | --- | --- |
| CYP2C19 | 141QK (reduced efflux transporter activity) | 1 | Use 1.25 times the standard dose. | N/A | Yes |
|  | 141KK (strongly reduced efflux transporter activity) | 2 | Use 1.4 times the standard dose. | N/A | Yes |
| HLA-B | *58:01 | 3 | Allopurinol is contraindicated. | Strong | Yes |

**Amikacin**

| **Gene** | **Genotype** | **#** | **Recommendation** | **Classification** | **Potentially actionable?** |
| --- | --- | --- | --- | --- | --- |
| MT-RNR1 | m.827A>G | 4 | Use aminoglycoside antibiotics at standard doses for the shortest feasible course with therapeutic dose monitoring. Evaluate regularly for hearing loss in line with local guidance. | Strong | Yes |

**Amitriptyline**

| **CYP2D6** | **CYP2C19** | **#** | **Recommendation** | **Classification** | **Potentially actionable?** |
| --- | --- | --- | --- | --- | --- |
| Poor metaboliser | Intermediate/normal metaboliser | 5 | Avoid amitriptyline use. If amitriptyline is warranted, consider a 50% reduction of recommended starting dose. Utilising therapeutic drug monitoring to guide dose adjustments is strongly recommended. | Optional | Yes |
|  | Rapid/ultrarapid metaboliser | 6 | Avoid amitriptyline use; If amitriptyline is warranted, utilise therapeutic drug monitoring to guide dose adjustment. Utilising therapeutic drug monitoring to guide dose adjustments is strongly recommended.** | Optional | Yes |
| Intermediate metaboliser | Poor metaboliser | 6 | Avoid amitriptyline use; If amitriptyline is warranted, utilise therapeutic drug monitoring to guide dose adjustment. Utilising therapeutic drug monitoring to guide dose adjustments is strongly recommended.** | Optional | Yes |
|  | Intermediate metaboliser | 7 | Consider a 25% reduction of recommended starting dose. Utilising therapeutic drug monitoring to guide dose adjustments is strongly recommended.** | Optional | Yes |
|  | Normal metaboliser | 7 | Consider a 25% reduction of recommended starting dose. Utilise therapeutic drug monitoring to guide dose adjustments.** | Moderate | Yes |
|  | Rapid/ultrarapid metaboliser | 8 | Consider alternative drug not metabolised by CYP2C19; If amitriptyline is warranted, utilise therapeutic drug monitoring to guide dose adjustment. Utilising therapeutic drug monitoring to guide dose adjustments is strongly recommended.* | Optional | Yes |
| Normal metaboliser | Poor metaboliser | 5 | Avoid amitriptyline use. If amitriptyline is warranted, consider a 50% reduction of recommended starting dose. Utilising therapeutic drug monitoring to guide dose adjustments is strongly recommended. | Optional | Yes |
|  | Intermediate metaboliser | 9 | Initiate therapy with recommended starting dose. | Strong | No |
|  | Rapid/ultrarapid metaboliser | 8 | Consider alternative drug not metabolised by CYP2C19; If amitriptyline is warranted, utilise therapeutic drug monitoring to guide dose adjustment.* | Optional | Yes |
| Ultrarapid metaboliser | Normal metaboliser | 10 | Avoid amitriptyline use; If amitriptyline is warranted, consider titrating to a higher target dose (compared to normal metabolisers). Utilise therapeutic drug monitoring to guide dose adjustment.** | Strong | Yes |
|  | Intermediate/rapid/ultrarapid metaboliser | 6 | Avoid amitriptyline use; If amitriptyline is warranted, utilise therapeutic drug monitoring to guide dose adjustment. Utilising therapeutic drug monitoring to guide dose adjustments is strongly recommended.** | Optional | Yes |

* Recommendations that have been combined for analysis due to similarity.

** Recommendations that have been combined with nortriptyline recommendations due to similarity.

**Nortriptyline**

| **Gene** | **Phenotype** | **#** | **Recommendation** | **Classification** | **Potentially actionable?** |
| --- | --- | --- | --- | --- | --- |
| CYP2D6 | Poor metaboliser | 6 | Avoid tricyclic use due to potential for side effects. Consider alternative drug not metabolised by CYP2D6. If a tricyclic antidepressant is warranted, consider a 50% reduction of recommended starting dose. Titrate dose to observed clinical response with symptom improvement and minimal (if any) side effects. Utilise therapeutic drug monitoring to guide dose adjustments.** | Strong | Yes |
|  | Intermediate metaboliser | 7 | Consider a 25% reduction of recommended starting dose. Titrate dose to observed clinical response with symptom improvement and minimal (if any) side effects. Utilise therapeutic drug monitoring to guide dose adjustments.** | Moderate | Yes |
|  | Ultrarapid metaboliser | 10 | Avoid tricyclic use due to potential lack of efficacy. Consider alternative drug not metabolised by CYP2D6. If a tricyclic antidepressant is warranted, consider titrating to a higher target dose (compared to normal metabolisers). Titrate dose to observed clinical response with symptom improvement and minimal (if any) side effects. Utilise therapeutic drug monitoring to guide dose adjustments.** | Strong | Yes |

** Recommendations that have been combined with amitriptyline recommendations due to similarity.

**Atazanavir**

| **Gene** | **Phenotype** | **#** | **Recommendation** | **Classification** | **Potentially actionable?** |
| --- | --- | --- | --- | --- | --- |
| UGT1A1 | Poor metaboliser | 11 | Consider an alternative agent particularly where jaundice would be of concern to the patient. If atazanavir is to be prescribed, there is a high likelihood of developing jaundice that will result in atazanavir discontinuation (at least 20% and as high as 60%). | Strong | Yes |
|  | Intermediate metaboliser | 12 | There is no need to avoid prescribing of atazanavir based on UGT1A1 genetic test result. Inform the patient that some patients stop atazanavir because of jaundice (yellow eyes and skin), but that this patient's genotype makes this unlikely (less than about a 1 in 20 chance of stopping atazanavir because of jaundice). | Strong | No |

**Azathioprine**

| **Gene** | **Phenotype** | **#** | **Recommendation** | **Classification** | **Potentially actionable?** |
| --- | --- | --- | --- | --- | --- |
| NUDT15 or TPMT | Poor metaboliser | 13 | For non-malignant conditions, consider alternative non-thiopurine immunosuppressant therapy. For malignant conditions, start with drastically reduced normal daily doses (reduce daily dose by 10-fold) and adjust doses of azathioprine based on degree of myelosuppression and disease-specific guidelines. Allow 4-6 weeks to reach steady-state after each dose adjustment (PMID 16530578, 11302950, 15606506, 16530532, 12477776). | Strong | Yes |
|  | Intermediate metaboliser | 14 | Start with reduced starting doses (30-80% of normal dose) if normal starting dose is 2-3mg/kg/day (e.g., 0.6-2.4mg/kg/day), and adjust doses of azathioprine based on degree of myelosuppression and disease-specific guidelines. Allow 2-4 weeks to reach steady-state after each dose adjustment (PMID 20354201, 11302950, 15606506, 16530532). | Strong | Yes |

**Capecitabine / Fluorouracil**

| **Gene** | **Phenotype** | **#** | **Recommendation** | **Classification** | **Potentially actionable?** |
| --- | --- | --- | --- | --- | --- |
| DPYD | Intermediate metaboliser | 15 | Reduce starting dose by 50% followed by titration of dose based on toxicity or therapeutic drug monitoring (if available). | Moderate | Yes |

**Carbamazepine**

| **Gene** | **Genotype** | **#** | **Recommendation** | **Classification** | **Potentially actionable?** |
| --- | --- | --- | --- | --- | --- |
| HLA-A | *31:01 | 16* | If patient is carbamazepine-naïve and alternative agents are available, do not use carbamazepine.* | Strong | Yes |
|  | *15:02 | 16* | If patient is carbamazepine-naïve, do not use carbamazepine.* | Strong | Yes |

**Citalopram / Escitalopram**

| **Gene** | **Phenotype** | **#** | **Recommendation** | **Classification** | **Potentially actionable?** |
| --- | --- | --- | --- | --- | --- |
| CYP2C19 | Poor metaboliser | 17 | Consider a clinically appropriate antidepressant not predominantly metabolised by CYP2C19. If citalopram or escitalopram are clinically appropriate, consider a lower starting dose, slower titration schedule and 50% reduction of the standard maintenance dose as compared to normal metabolisers. | Strong | Yes |
|  | Intermediate metaboliser | 18 | Initiate therapy with recommended starting dose. Consider a slower titration schedule and lower maintenance dose than normal metabolisers. | Moderate | Yes |
|  | Rapid metaboliser | 19 | Initiate therapy with recommended starting dose. If patient does not adequately respond to recommended maintenance dosing, consider titrating to a higher maintenance dose or switching to a clinically appropriate alternative antidepressant not predominantly metabolized by CYP2C19. | Optional | Yes, if trialed with inadequate response |
|  | Ultrarapid metaboliser | 20 | Consider a clinically appropriate alternative antidepressant not predominantly metabolised by CYP2C19. If citalopram or escitalopram are clinically appropriate, and adequate efficacy is not achieved at standard maintenance dosing, consider titrating to a higher maintenance dose. | Strong | Yes |

**Codeine / Tramadol**

| **Gene** | **Phenotype** | **#** | **Recommendation** | **Classification** | **Potentially actionable?** |
| --- | --- | --- | --- | --- | --- |
| CYP2D6 | Poor metaboliser | 21 | Use codeine/tramadol label recommended age- or weight-specific dosing. If no response and opioid use is warranted, consider non-tramadol/codeine opioid. | Optional | Yes, if trialed with inadequate response |
|  | Intermediate metaboliser | 22 | Avoid codeine/tramadol use because of possibility of diminished analgesia. If opioid use is warranted, consider a non-tramadol/codeine opioid. | Strong | Yes |

**Flecainide**

| **Gene** | **Phenotype** | **#** | **Recommendation** | **Classification** | **Potentially actionable?** |
| --- | --- | --- | --- | --- | --- |
| CYP2D6 | Poor metaboliser | 23 | Reduce the dose to 50% of the standard dose and record an ECG and monitor the plasma concentration. | N/A | Yes |
|  | Intermediate metaboliser | 24 | Indications other than diagnosis of Brugada syndrome: Reduce the dose to 75% of the standard dose and record an ECG and monitor the plasma concentration.  Provocation test for diagnosis of Brugada syndrome: No action required.  At a dose of 2 mg/kg body weight to a maximum of 150mg, the response is better for patients with alleles that result in reduced activity. All 5 patients with these alleles and 20% of the patients with two fully active alleles exhibited a response within 30 minutes. | N/A | Yes, if used for indications other than Brugada syndrome |
|  | Ultrarapid metaboliser | 25 | There are no data about the pharmacokinetics and/ or the effects of flecainide in ultrarapid metabolisers. 1. Monitor the plasma concentration as a precaution and record an ECG or select an alternative. Examples of anti-arrhythmic drugs that are not metabolised via CYP2D6 ( or to a lesser extent) include sotalol, disopyramide, quinidine and amiodarone. | N/A | Yes |

**Flucloxacillin**

| **Gene** | **Genotype** | **#** | **Recommendation** | **Classification** | **Potentially actionable?** |
| --- | --- | --- | --- | --- | --- |
| HLA-B | *57:01 | 26 | Regularly monitor the patient's liver function and choose an alternative if liver enzymes and/or bilirubin levels are elevated. | N/A | Yes |

**Fluvoxamine**

| **Gene** | **Phenotype** | **#** | **Recommendation** | **Classification** | **Potentially actionable?** |
| --- | --- | --- | --- | --- | --- |
| CYP2D6 | Poor metaboliser | 27 | Consider a 25-50% lower starting dose and slower titration schedule as compared to normal metabolisers or consider a clinically appropriate alternative antidepressant not predominantly metabolised by CYP2D6. | Optional | Yes |
|  | Intermediate metaboliser | 28 | Initiate therapy with recommended starting dose. | Moderate | No |
|  | Ultrarapid metaboliser | 29 | No recommendation due to lack of evidence. | N/A | No |

**Irinotecan**

| **Gene** | **Phenotype** | **#** | **Recommendation** | **Classification** | **Potentially actionable?** |
| --- | --- | --- | --- | --- | --- |
| UGT1A1 | Poor metaboliser | 30 | Start with 70% of the standard dose. If the patient tolerates this initial dose, the dose can be increased, guided by the neutrophil count. | N/A | Yes |

**Mercaptopurine**

| **Gene** | **Phenotype** | **#** | **Recommendation** | **Classification** | **Potentially actionable?** |
| --- | --- | --- | --- | --- | --- |
| NUDT15 | Poor metaboliser | 31 | For malignancy, initiate dose at 10mg/m^2^/day and adjust dose based on myelosuppression and disease-specific guidelines. Allow 4-6 weeks to reach steady state after each dose adjustment. If myelosuppression occurs, emphasis should be on reducing mercaptopurine over other agents. For non-malignant conditions, consider alternative non-thiopurine immunosuppressant therapy (PMID 20354201, 1960624, 11302950, 16530532). | Strong | Yes |
|  | Intermediate metaboliser | 32 | Start with reduced starting doses (30-80% of normal dose) if normal starting dose is ≥75mg/m2/day or ≥1.5mg/kg/day (e.g., start at 22.5-60mg/m^2^/day or 0.45-1.2mg/kg/day) and adjust doses of mercaptopurine based on degree of myelosuppression and disease-specific guidelines. Allow 2-4 weeks to reach steady-state after each dose adjustment. If myelosuppression occurs, and depending on other therapy, emphasis should be on reducing mercaptopurine over other agents (PMID 20354201, 18685564, 8857546, 18987654, 20010622, 16401827, 11302950, 16530532, 9634537). If normal starting dose is already <75mg/m^2^/day or <1.5 mg/kg/day, dose reduction may not be recommended. | Strong | Yes |
| TPMT | Poor metaboliser | 33 | For malignancy, start with drastically reduced doses (reduce daily dose by 10-fold and reduce frequency to thrice weekly instead of daily, e.g., 10 mg/m^2^/day given just 3 days/week) and adjust doses of mercaptopurine based on degree of myelosuppression and disease-specific guidelines. Allow 4-6 weeks to reach steady-state after each dose adjustment. If myelosuppression occurs, emphasis should be on reducing mercaptopurine over other agents. For non-malignant conditions, consider alternative non-thiopurine immunosuppressant therapy (PMID 20354201, 1960624, 11302950, 16530532). | Strong | Yes |
|  | Intermediate metaboliser | 32 | Start with reduced starting doses (30-80% of normal dose) if normal starting dose is ≥75mg/m2/day or ≥1.5mg/kg/day (e.g., start at 22.5-60mg/m^2^/day or 0.45-1.2mg/kg/day) and adjust doses of mercaptopurine based on degree of myelosuppression and disease-specific guidelines. Allow 2-4 weeks to reach steady-state after each dose adjustment. If myelosuppression occurs, and depending on other therapy, emphasis should be on reducing mercaptopurine over other agents (PMID 20354201, 18685564, 8857546, 18987654, 20010622, 16401827, 11302950, 16530532, 9634537). If normal starting dose is already <75mg/m^2^/day or <1.5 mg/kg/day, dose reduction may not be recommended. | Strong | Yes |

**Methylene Blue**

| **Gene** | **Phenotype** | **#** | **Recommendation** | **Classification** | **Potentially actionable?** |
| --- | --- | --- | --- | --- | --- |
| G6PD | Deficiency | 34 | Avoid use. | Strong | Yes |
|  | Variable | 35 | To ascertain G6PD status, enzyme activity must be measured. Drug use should be guided as per the recommendations based on the activity-based phenotype. | Moderate | No |

**Omeprazole / Pantoprazole**

| **Gene** | **Phenotype** | **#** | **Recommendation** | **Classification** | **Potentially actionable?** |
| --- | --- | --- | --- | --- | --- |
| CYP2C19 | Poor/intermediate metaboliser | 36 | Initiate standard starting daily dose. For chronic therapy (>12 weeks) and efficacy achieved, consider 50% reduction in daily dose and monitor for continued efficacy. | Moderate/ optional | Yes, if used for >12 weeks |
|  | Rapid metaboliser | 37 | Initiate standard starting daily dose. Consider increasing dose by 50-100% for the treatment of H. pylori infection and erosive esophagitis. Daily dose may be given in divided doses. Monitor for efficacy. | Moderate | Yes |
|  | Ultrarapid metaboliser | 38 | Increase starting daily dose by 100%. Daily dose may be given in divided doses. Monitor for efficacy. | Optional | Yes |

**Ondansetron / Tropisetron**

| **Gene** | **Phenotype** | **#** | **Recommendation** | **Classification** | **Potentially actionable?** |
| --- | --- | --- | --- | --- | --- |
| CYP2D6 | Poor/intermediate metaboliser | 39 | Insufficient evidence demonstrating clinical impact based on CYP2D6 genotype. Initiate therapy with recommended starting dose. | N/A | No |
|  | Ultrarapid metaboliser | 40 | Select alternative drug not predominantly metabolised by CYP2D6 (e.g. granisetron). | Moderate | Yes |

**Paroxetine**

| **Gene** | **Phenotype** | **#** | **Recommendation** | **Classification** | **Potentially actionable?** |
| --- | --- | --- | --- | --- | --- |
| CYP2D6 | Poor metaboliser | 41 | Consider a 50% reduction in recommended starting dose, slower titration schedule, and a 50% lower maintenance dose as compared to normal metabolisers. | Moderate | Yes |
|  | Intermediate metaboliser | 42 | Initiate therapy with recommended starting dose.*** | Moderate | No |
|  | Intermediate metaboliser | 43 | Consider a lower starting dose and slower titration schedule as compared to normal metabolisers. | Optional | Yes |
|  | Ultrarapid metaboliser | 44 | Select alternative drug not predominantly metabolised by CYP2D6. | Moderate | Yes |

*** Recommendation provided for small number of patients before being updated.

**Rasburicase**

| **Gene** | **Phenotype** | **#** | **Recommendation** | **Classification** | **Potentially actionable?** |
| --- | --- | --- | --- | --- | --- |
| G6PD | Deficiency | 45 | Avoid use. | Strong | Yes |
|  | Variable | 46 | To ascertain G6PD status, enzyme activity must be measured. Drug use should be guided as per the recommendations based on the activity-based phenotype. | Moderate | No |

**Sertraline**

| **CYP2B6** | **CYP2C19** | **#** | **Recommendation** | **Classification** | **Potentially actionable?** |
| --- | --- | --- | --- | --- | --- |
| Poor metaboliser | Poor metaboliser | 47 | Select an alternative antidepressant not primarily metabolised by CYP2C19 or CYP2B6. | Optional | Yes |
|  | Intermediate metaboliser | 48 | Consider a lower starting dose, slower titration schedule and 50% reduction of standard maintenance dose as compared to CYP2B6 normal metabolisers. | Optional | Yes |
|  | Normal metaboliser | 49 | Consider a lower starting dose, slower titration schedule and 50% reduction of standard maintenance dose as compared to CYP2B6 normal metabolisers or select a clinically appropriate alternative antidepressant not predominantly metabolised by CYP2B6.* | Optional | Yes |
|  | Rapid/ultrarapid metaboliser | 50 | Initiate therapy with recommended starting dose. | Optional | No |
| Intermediate metaboliser | Poor metaboliser | 49 | Consider a lower starting dose, slower titration schedule and 50% reduction of standard maintenance dose as compared to CYP2C19 normal metabolisers or select a clinically appropriate alternative antidepressant not predominantly metabolised by CYP2C19.* | Moderate | Yes |
|  | Intermediate/normal metaboliser | 51 | Initiate therapy with recommended starting dose. Consider a slower titration schedule and lower maintenance dose than normal metabolisers. | Optional/moderate | Yes |
|  | Rapid/ultrarapid metaboliser | 50 | Initiate therapy with recommended starting dose. | Moderate | No |
| Rapid metaboliser | Intermediate metaboliser | 50 | Initiate therapy with recommended starting dose. | Moderate | No |
| Normal metaboliser | Poor metaboliser | 49 | Consider a lower starting dose, slower titration schedule and 50% reduction of standard maintenance dose as compared to CYP2C19 normal metabolisers or select a clinically appropriate alternative antidepressant not predominantly metabolised by CYP2C19.* | Moderate | Yes |
|  | Intermediate metaboliser | 51 | Initiate therapy with recommended starting dose. Consider a slower titration schedule and lower maintenance dose than normal metabolisers. | Moderate | Yes |
|  | Rapid/ultrarapid metaboliser | 50 | Initiate therapy with recommended starting dose. | Strong | No |

* Recommendations that were combined due to similarity.

**Simvastatin**

| **Gene** | **Phenotype** | **#** | **Recommendation** | **Classification** | **Potentially actionable?** |
| --- | --- | --- | --- | --- | --- |
| SLCO1B1 | Decreased/poor function | 52 | Prescribe an alternative statin depending on the desired potency (see Figure 1 of PMID: 35152405 for recommendations for alternative statins). If simvastatin therapy is warranted, limit dose to <20mg/day. | Strong | Yes |
|  | Increased function | 53 | Prescribe desired starting dose and adjust doses based on disease-specific guidelines. | Strong | No |

**Tacrolimus**

| **Gene** | **Phenotype** | **#** | **Recommendation** | **Classification** | **Potentially actionable?** |
| --- | --- | --- | --- | --- | --- |
| CYP3A5 | Intermediate/extensive/normal metaboliser | 54 | Increase starting dose 1.5 to 2 times recommended starting dose. Total starting dose should not exceed 0.3 mg/kg/day. Use therapeutic drug monitoring to guide dose adjustments. | Strong | Yes |

**Thioguanine**

| **Gene** | **Phenotype** | **#** | **Recommendation** | **Classification** | **Potentially actionable?** |
| --- | --- | --- | --- | --- | --- |
| NUDT15 | Poor metaboliser | 55 | Reduce starting doses to 25% of normal dose and adjust doses of thioguanine based on degree of myelosuppression and disease-specific guidelines. Allow 4-6 weeks to reach steady-state after each dose adjustment. In setting of myelosuppression, emphasis should be on reducing thioguanine over other agents. For non-malignant conditions, consider alternative non-thiopurine immunosuppressant therapy (PMID 20354201). | Strong | Yes |
|  | Intermediate metaboliser | 56 | Start with reduced doses (50% to 80% of normal dose) if normal starting dose is ≥40-60 mg/m2/day (e.g., 20-48mg/m^2^/day) and adjust doses of thioguanine based on degree of myelosuppression and disease-specific guidelines. Allow 2-4 weeks to reach steady-state after each dose adjustment. If myelosuppression occurs, and depending on other therapy, emphasis should be on reducing thioguanine over other agents (PMID 20354201, 11037857). | Moderate | Yes |
| TPMT | Poor metaboliser | 57 | Start with drastically reduced doses (PMID 11037857) (reduce daily dose by 10-fold and dose thrice weekly instead of daily) and adjust doses of thioguanine based on degree of myelosuppression and disease-specific guidelines. Allow 4-6 weeks to reach steady-state after each dose adjustment. If myelosuppression occurs, emphasis should be on reducing thioguanine over other agents. For non-malignant conditions, consider alternative non-thiopurine immunosuppressant therapy (PMID 20354201 ). | Strong | Yes |
|  | Intermediate metaboliser | 56 | Start with reduced doses (50% to 80% of normal dose) if normal starting dose is ≥40-60 mg/m2/day (e.g., 20-48mg/m^2^/day) and adjust doses of thioguanine based on degree of myelosuppression and disease-specific guidelines. Allow 2-4 weeks to reach steady-state after each dose adjustment. If myelosuppression occurs, and depending on other therapy, emphasis should be on reducing thioguanine over other agents (PMID 20354201, 11037857). | Moderate | Yes |

**Voriconazole**

| **Gene** | **Phenotype** | **#** | **Recommendation** | **Classification** | **Potentially actionable?** |
| --- | --- | --- | --- | --- | --- |
| CYP2C19 | Poor metaboliser | 58 | Choose an alternative agent that is not dependent on CYP2C19 metabolism as primary therapy in lieu of voriconazole. Such agents include liposomal amphotericin B and posaconazole. In the event that voriconazole is considered to be the most appropriate agent, based on clinical advice, for a patient with poor metaboliser genotype, voriconazole should be administered at a preferably lower than standard dosage with careful therapeutic drug monitoring.* | Moderate | Yes |
|  | Intermediate metaboliser | 59 | Initiate therapy with recommended standard of care dosing.* | Moderate | No |
|  | Rapid metaboliser | 60 | Initiate therapy with recommended standard of care dosing. Use therapeutic drug monitoring to titrate dose to therapeutic trough concentrations.* | Moderate | Yes |
|  | Ultrarapid metaboliser | 58 | Choose an alternative agent that is not dependent on CYP2C19 metabolism as primary therapy in lieu of voriconazole. Such agents include liposomal amphotericin B and posaconazole.* | Moderate | Yes |
