## Supplementary Table S2: Prescribing events and adherence rates/grades by individual PGx recommendation for "Adherence to International Pharmacogenomic Recommendations in Paediatric Cancer Care: A Cohort Analysis Embedded Within the MARVEL-PIC Randomised Trial"

| Drug | Rec # | n (patients) | n (events) | Explicitly Followed | Inadvertently Followed | Not Followed | Rate (Explicit Adherence) | Grade (Explicit Adherence) | Rate (Overall Adherence) | Grade (Overall Adherence) |
| --- | --- | --- | --- | --- | --- | --- | --- | --- | --- | --- |
| Amitriptyline | 5 | 1 | 3 | 0 | 3 | 0 | 0.0% | Very low | 100.0% | High |
|  | 8 | 1 | 9 | 0 | 9 | 0 | 0.0% | Very low | 100.0% | High |
| Amitriptyline / Nortriptyline | 6 | 9 | 36 | 0 | 36 | 0 | 0.0% | Very low | 100.0% | High |
| Carbamazepine | 16 | 1 | 4 | 0 | 4 | 0 | 0.0% | Very low | 100.0% | High |
| Codeine / Tramadol | 21 | 1 | 1 | 0 | 1 | 0 | 0.0% | Very low | 100.0% | High |
|  | 22 | 11 | 72 | 0 | 72 | 0 | 0.0% | Very low | 100.0% | High |
| Mercaptopurine | 31 | 1 | 3 | 3 | 0 | 0 | 100.0% | High | 100.0% | High |
|  | 32 | 3 | 12 | 10 | 0 | 2 | 83.3% | High | 83.3% | High |
|  | 33 | 1 | 1 | 1 | 0 | 0 | 100.0% | High | 100.0% | High |
| Omeprazole / Pantoprazole | 36 | 1 | 5 | 0 | 0 | 5 | 0.0% | Very low | 0.0% | Very low |
|  | 37 | 17 | 36 | 9 | 0 | 27 | 25.0% | Low | 25.0% | Low |
|  | 38 | 1 | 1 | 0 | 0 | 1 | 0.0% | Very low | 0.0% | Very low |
| Ondansetron / Tropisetron | 40 | 6 | 69 | 21 | 3 | 45 | 30.4% | Low | 34.8% | Low |
| Sertraline | 51 | 1 | 5 | 0 | 0 | 0 | 0.0% | Very low | 0.0% | Very low |
| Thioguanine | 57 | 1 | 2 | 1 | 0 | 1 | 50.0% | Moderate | 50.0% | Moderate |
| Voriconazole | 58 | 3 | 14 | 0 | 14 | 0 | 0.0% | Very low | 100.0% | High |
|  | 60 | 5 | 15 | 12 | 3 | 0 | 80.0% | High | 100.0% | High |

**Supplementary Table S2:** Prescribing events and adherence rates/grades by individual PGx recommendation
