## Supplemental Data 1 for "Adherence to International Pharmacogenomic Recommendations in Paediatric Cancer Care: A Cohort Analysis Embedded Within the MARVEL-PIC Randomised Trial"

|  | Explicit adherence | Cramér’s V | Overall adherence | Cramér’s V |
| --- | --- | --- | --- | --- |
| Disease group  (liquid vs solid vs brain vs transplant) | p=0.1866 | 0.3030 | p=0.628 | 0.235 |
| Age group  (<5 vs 5-9.9 vs ≥10) | p=0.8689 | 0.1764 | p=0.3949 | 0.2532 |
| Drug type  (supportive vs chemotherapy vs treatment) | p=0.181 | 0.7071 | p=0.3714 | 0.6614 |
| Recommendation strength  (optional vs moderate vs strong) | p=0.1456 | 0.5072 | p=0.2449 | 0.5089 |

**Supplementary Table S3:** Fisher-Freeman-Halton p-values and Cramér’s V effect sizes for different patient/drug/recommendation subgroups
